# A deep learning approach for cancer diagnosis exclusively from raw sequencing fragments of bisulfite-treated plasma cell-free DNA

**DOI:** 10.1101/2023.08.08.23293813

**Authors:** Jilei Liu, Hongru Shen, Jiani Hu, Xilin Shen, Chao Zhang, Yichen Yang, Meng Yang, Wei Wang, Yang Li, Qiang Zhang, Jilong Yang, Kexin Chen, Xiangchun Li

## Abstract

Early cancer diagnosis from bisulfite-treated cell-free DNA (cfDNA) fragments require tedious data analytical procedures. Here, we present a Deep-learning-based approach for Early Cancer Interception and DIAgnosis (DECIDIA) that can achieve accurate cancer diagnosis exclusively from bisulfite-treated cfDNA sequencing fragments. DECIDIA relies on feature representation learning of DNA fragments and weakly supervised learning for classification. We systematically evaluate the performance of DECIDIA for cancer diagnosis and cancer-type prediction on a curated dataset of 5389 samples that consist of colorectal cancer (CRC, n = 1574), hepatocellular cell carcinoma (HCC, n = 1181), lung cancer (n = 654) and non-cancer control (n=1980). DECIDIA achieves an area under the receiver operating curve (AUROC) of 0.980 (95% CI, 0.976-0.984) in ten-fold cross validation settings on the CRC dataset, outperforming benchmarked methods that are based on fragmentation profiles and methylation intensities. Noticeably, DECIDIA achieves an AUROC of 0.910 (95% CI, 0.896-0.924) on the externally independent HCC testing set although there was no HCC data used in model development. In the settings of cancer-type classification, we observed that DECIDIA achieves a micro-average AUROC of 0.963 (95% CI, 0.960-0.966) and an overall accuracy of 82.8% (95%CI, 81.8% - 83.9%). In addition, we distilled four sequence signatures from the raw sequencing reads that exhibited differential patterns in cancer versus control and among different cancer types. Our approach represents a new paradigm towards eliminating the tedious data analytical procedures for liquid biopsy that uses bisulfite-treated cfDNA methylome.

## Introduction

Plasma cell-free DNA (cfDNA) testing has been widely studied for early cancer diagnosis.(Chabon et al., 2020) cfDNA is mainly released from cell apoptosis, necrosis and active secretion.(Jahr et al., 2001; Thakur et al., 2014) The half-life of cfDNA in blood is estimated to be ranging from 16 minutes to 2.5 hours.(Diehl et al., 2008; To et al., 2003) Therefore, cfDNA is seen as a real-time snapshot manifesting physiological condition of the human body, offering diagnostic information. We have been witnessing wide application of cfDNA in multiple clinical settings besides early cancer diagnosis.(Berchuck et al., 2022; Chan et al., 2022; Siravegna and Bardelli, 2014) Different modalities of cfDNA have been investigated for cancer diagnosis including mutations(Forshew et al., 2012; Kinde et al., 2011), fragment patterns(Cristiano et al., 2019; X. Zhang et al., 2022; Zhou et al., 2022), and methylation profiles(Chemi et al., 2022; Klein et al., 2021; Liang et al., 2021; Xu et al., 2017). The methylome of bisulfite-treated cfDNA fragments are attracting the most attentions among the other modalities.(Chen et al., 2020; Guo et al., 2017; Kang et al., 2017; Li et al., 2018; Liang et al., 2021; Liu et al., 2020; Shen et al., 2018; Xu et al., 2017)

DNA methylation is an earlier event in tumorigenesis. Besides, DNA methylation patterns are tissue-specific.(Moss et al., 2018) Bisulfite sequencing enables single-base resolution for characterizing DNA methylation. The core idea of bisulfite sequencing is that bisulfite treatment can deaminate unmethylated cytosine to uracil while methylated cytosine remains unchanged. Profiling plasma cfDNA by bisulfite sequencing is a promising solution for developing blood-based early cancer diagnosis.(Feng et al., 2019) The Circulating Cell-free Genome Atlas (CCGA) project is one of the representative endeavors devoted to developing blood-based early cancer diagnosis by profiling plasm cfDNA with bisulfite sequencing. Three studies from the CCGA group reported that the methylation patterns of plasma cfDNA are able to precisely detect multiple cancer types at earlier stage.(Klein et al., 2021; Liu et al., 2020, 2018) Meanwhile, Xu and colleagues also reported high sensitivity and specificity in the diagnosis of colorectal cancer and hepatocellular carcinoma by profiling cfDNA methylation via targeted-bisulfite sequencing.(Luo et al., 2020; Xu et al., 2017) However, all of these studies rely upon quantification of cfDNA methylation abundance which requires base conversions and sequence alignment in advance.(Luo et al., 2021)

Transformer-based pre-trained language models achieve excellent performance in many natural language understanding such as translation and semantic analysis.(Devlin et al., n.d.; Radford et al., n.d.; Vaswani et al., 2017) This solution has also been adopted in learning the feature representation of DNA and protein sequences. For example, deep language models pre-trained on human reference genome can capture the global and transferable representation of the genomic DNA sequence and achieve state-of-the-art performance at characterization of promoters, splice sites, and transcription factor binding sites.(Ji et al., n.d.) Multiple-instance learning is a type of weakly-supervised deep learning algorithm(Ilse et al., 2018; Lu et al., 2021a, 2021b) that is useful for classification in the scenario where only label of the individual is available but the labels of the instances of that individual are not available. By coupling with the attention-based mechanism, this weakly-supervised learning approach is able to identify the instances that are most relevant for classification.(Amores, 2013; Ilse et al., 2018)

In this study, we present a <u>d</u>eep-learning-based approach for <u>e</u>arly <u>c</u>ancer interception and <u>dia</u>gnosis called DECIDIA exclusively from the raw sequencing reads of bisulfite-treated plasma cfDNA fragments. DECIDIA consists of development of a transformer-based module for learning the features of DNA sequences and application of multiple-instance learning for aggregating multiple sequencing reads for classification. We systematically evaluated the performance of DECIDIA for cancer diagnosis and cancer type prediction on a curated dataset of 5389 samples that consist of colorectal cancer (CRC, n = 1574), hepatocellular cell carcinoma (HCC, n = 2140), lung cancer (n = 654) and non-cancer control (n=1980) subjected to targeted-bisulfite sequencing of plasma cfDNA. We demonstrated that DECIDIA is robust and efficient for varying number of sample size and sequencing reads. DECIDIA is quite simple in essence in that it directly uses raw sequencing reads instead of methylation abundance. DECIDIA eliminates the tedious data analytical procedures required in developing early cancer diagnosis tools using plasma cfDNA methylation. It will facilitate the development of cfDNA-based diagnostic tools.

## Results

### Overview of DECIDIA

Fundamentally, DECIDIA consists of two components including the development of a feature representation learning model for sequencing reads and construction of an attention-based multiple-instance learning model for sample-level classification by aggregating multiple reads from that sample. We employed the autoregressive language model(Radford et al., n.d.; S. Zhang et al., 2022) for learning DNA sequence feature. For an input DNA sequence ***S*** = {*b*_1_, *b*_2_, *b*_3_ …, *b*_*n*_}, the purpose of autoregressive modeling is to predict the base *b_i_* in the context of all its preceding bases, i.e., *b*_1_, *b*_2_, *b*_3_ …, *b_i_*_-1_. Data used to train the autoregressive model is the DNA sequences clipped equally from the human reference genome. We used the standard language modeling objective *L*(***S***) = ∑*_i_ log P*(*b_i_*|*b*_1_, *b*_2_, *b*_3_ …, *b_i_*_-1_; ***θ***) to maximize the likelihood.(Radford et al., n.d.) ***θ*** are the parameters of the autoregressive model that is used to model the conditional probability (See Methods). The trained autoregressive model was used to extract features for sequencing reads. Subsequently, the extracted features from sequencing reads obtained from each individual were fed into multiple-instance learning classifier. The attention-based multiple-instance classifier quantifies the influence of all instances from that individual on the classification label. Here, an instance refers to a sequencing read from that individual. Higher attention score indicates such an instance has greater discriminative capability. We trained this classifier iteratively by using cross-entropy loss. A flow diagram describing these procedures is shown in **Figure 1**.

**Figure 1.**
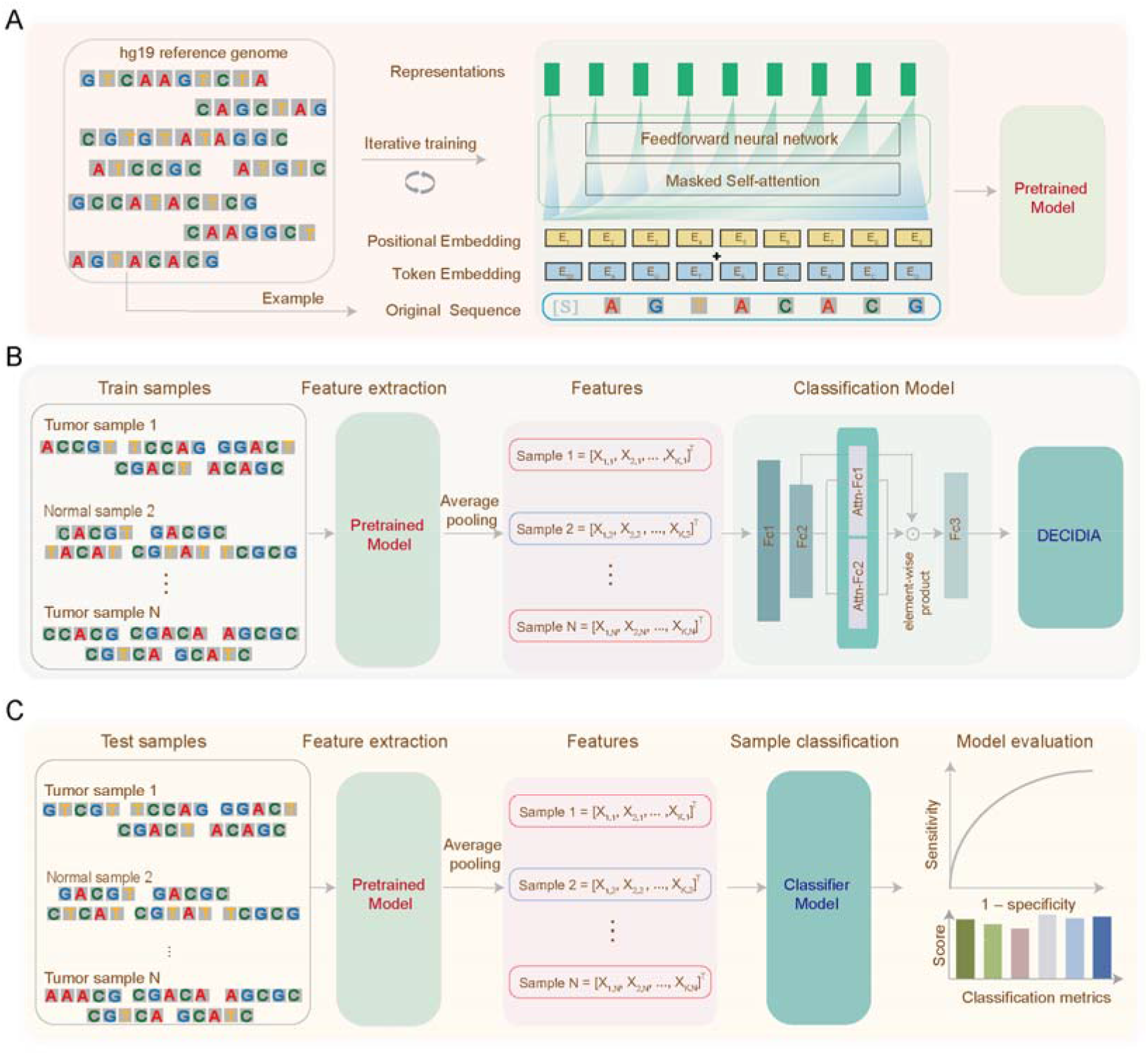
A flowchart depicting the development and evaluation of DECIDIA. (**A**) Development of an autoregressive language model on human reference genome for learning feature representations of DNA sequences. (**B**) Development of a multiple instance learning classifier with attention mechanism for cancer diagnosis and cancer type classification. **(C)** Performance evaluation on testing sets.

### High performance of DECIDIA in diagnosis of cancer

We employed ten-fold Monte Carlo cross-validation(Xu and Liang, 2001) to evaluate the cancer diagnostic performance of DECIDIA. For each cross-validation fold, we randomly divided the CRC dataset into training (n = 1000), validation (n = 200) and testing (n = 622) sets. Besides, we varied the number of training samples from 200 to 1000 and sequencing reads from 200 to 1000 for each cross-validation fold. We trained DECIDIA on the training set and monitor its performance with validation set at the end of each training epoch. We reported the classification performance of DECIDIA on the testing set with the checkpoint that performed the best on the validation set. We observed that DECIDIA achieved equally good performance with varying number of training samples and sequencing reads (**Figure 2E**). For example, DECIDIA trained with 1000 samples and 600 reads achieved an average AUROC of 0.980 (95% CI, 0.976-0.984) (**Figure 2A**), accuracy of 0.938 (95% CI,0.932-0.944), sensitivity of 0.972 (95% CI, 0.959 – 0.985) and specificity of 0.911(95% CI, 0.894 – 0.928) (**Figure 2B**); whereas DECIDIA trained with the minimum number of 200 training samples and 200 reads achieved an average AUROC of 0.939 (95%CI, 0.922 – 0.955), accuracy of 0.899 (95% CI, 0.870 – 0.927), sensitivity of 0.944 (95% CI, 0.910 – 0.977) and specificity of 0.862 (95% CI, 0.828 – 0.896). The performance of DECIDIA was increasing with more training samples and number of reads used. We observed that cancer patients and control individuals are distinguishable in the scatter plot generated from principle component analysis (PCA) of the features learned by DECIDIA (**Figure 2D**). The other classification metrics such as positive predictive rate, negative predictive rate and F1 metric were provided in **Supplementary Table S2**. DECIDIA outperformed fragmentation-based methods such as motif diversity score (AUROC = 0.557) and NMF-based deconvolution of fragmentation profile (AUROC = 0.66). Besides, DECIDIA also achieved better than machine learning method utializing methylation profile reported by Luo and colleagues (AUROC = 0.96)(Luo et al., 2020) (**Supplementary Figure 1**).

**Figure 2.**
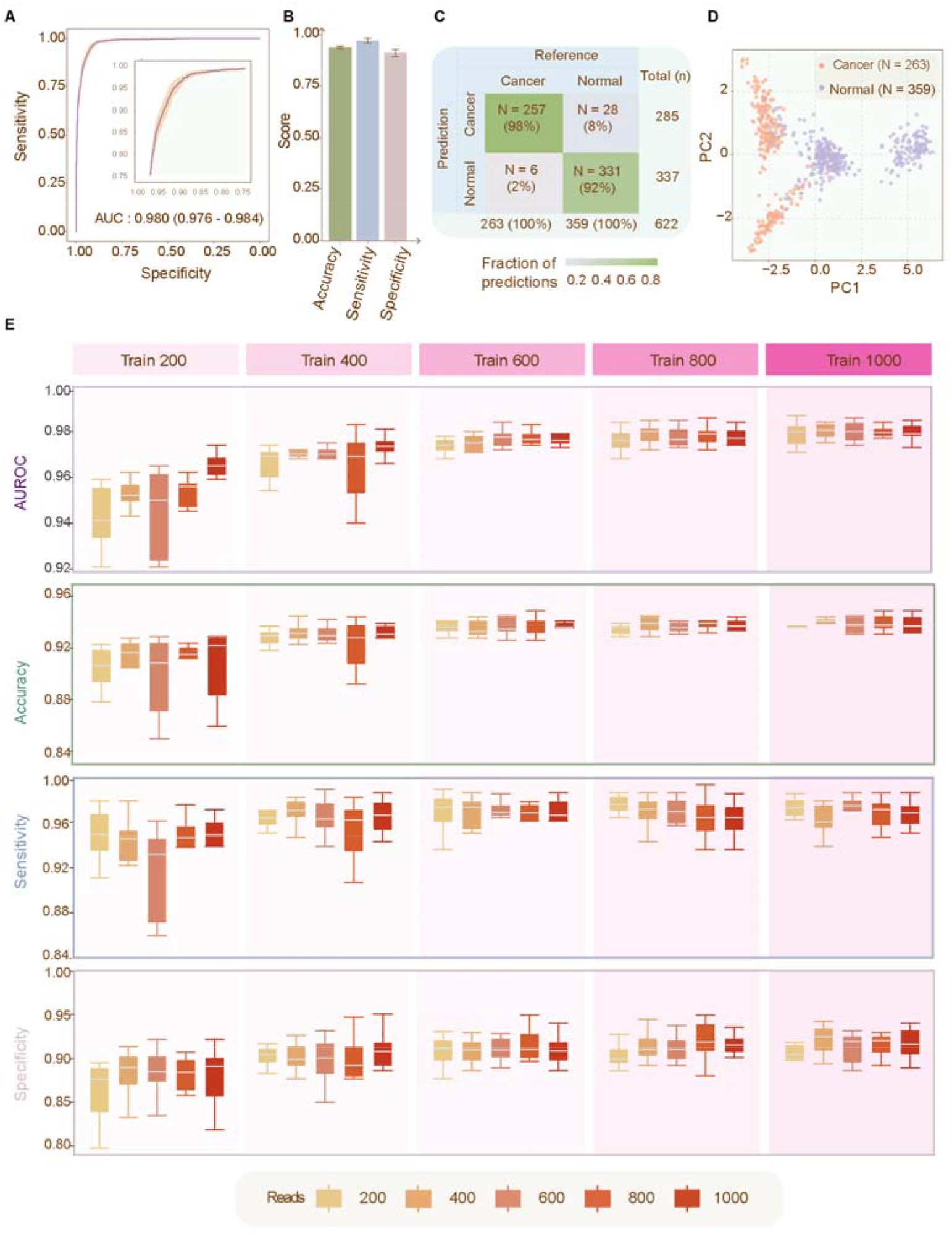
The classification performance of DECICIA in the diagnosis of cancer. **(A)**The ROC curve averaged from 10-fold Monte Carlo cross-validations. Insets: zoomed-in view of the ROC curve. **(B)** Barplot representation of accuracy, sensitivity and specificity. **(C)** Confusion matrix. **(D)** Scatter plot representation of the PCA of the learned features. **(E)** Boxplot representation of AUROC, accuracy, sensitivity and specificity for varying number of training samples and sequencing reads.

Noticeably, we observed that DECIDIA achieved an AUROC of 0.910 (95%CI, 0.896-0.924) on the externally independent hepatocellular cell carcinoma (HCC) testing set (n = 2140) even though no HCC data was involved in model training. The HCC testing set consists of 1181 patients with cancer and 959 controls. The ROC curve was shown in **Figure 3A**. The accuracy, sensitivity and specificity achieved by DECIDIA on this HCC testing set were 0.882 (95%CI, 0.868-0.896), 0.899 (95% CI, 0.881-0.916) and 0.861 (95% CI, 0.838-0.883), respectively (**Figure 3C**). Meanwhile, the PCA result from the extracted features for the HCC testing set are distinctive between HCC patients and controls (**Figure 3D)**.

**Figure 3.**
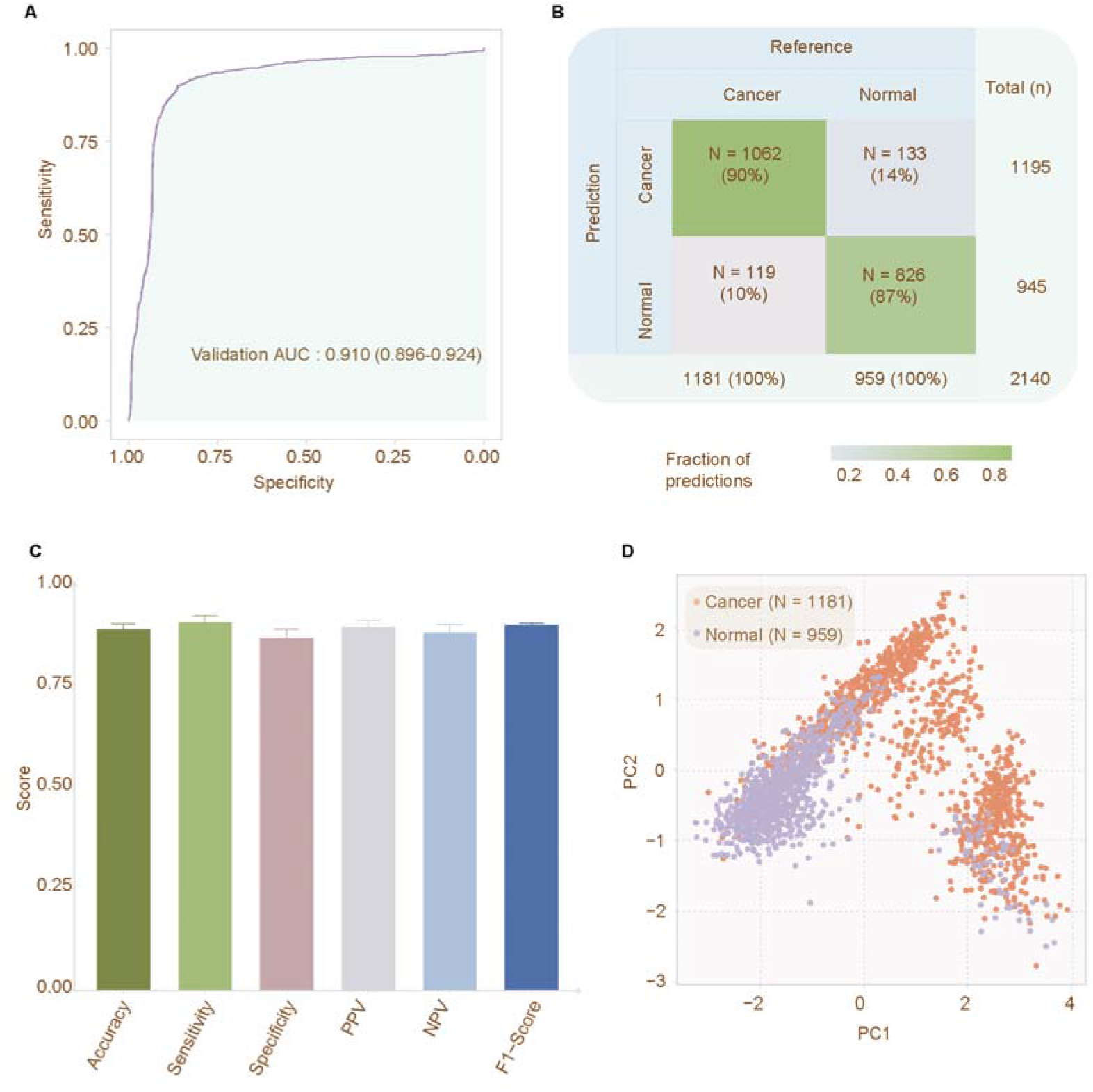
The classification performance of DECIDIA on the external HCC testing set. **(A)** The ROC curve. **(B)** Confusion matrix. **(C)** Barplot representation of accuracy, sensitivity, specificity, positive predictive value, negative predictive value and F1 score. **(D)** Scatter plot representation of the PCA of the learned features..

### High performance of DECIDIA in distinguishing different cancer types

We curated a total number of 3409 samples for the raw targeted-bisulfite sequencing data of plasma cfDNA from colorectal cancer (CRC, n = 1154), HCC (n = 1181) and lung cancer (n = 654) to evaluate the performance of DECIDIA on the detection of different cancer types. We randomly split this curated dataset into training (n = 2308), validation (n = 300) and testing (n = 800) sets. We used the training set to train DECIDIA and validation set to choose the best checkpoint and subsequently evaluated its performance on the testing set. On the testing set, DECIDIA achieved a micro-averaged AUROC of 0.963 (95% CI, 0.960-0.966) and an overall accuracy of 82.8% (95%CI, 81.8% - 83.9%) (**Figure 4B and Supplementary Table S3**). Different cancer types are visually distinguishable on the scatter plot generated from the PCA of the features extracted by DECIDIA (**Figure 4C**). The other classification metrics such as sensitivity, specificity and F1-score were shown in **Figure 4E and Supplementary Table S4**.

**Figure 4.**
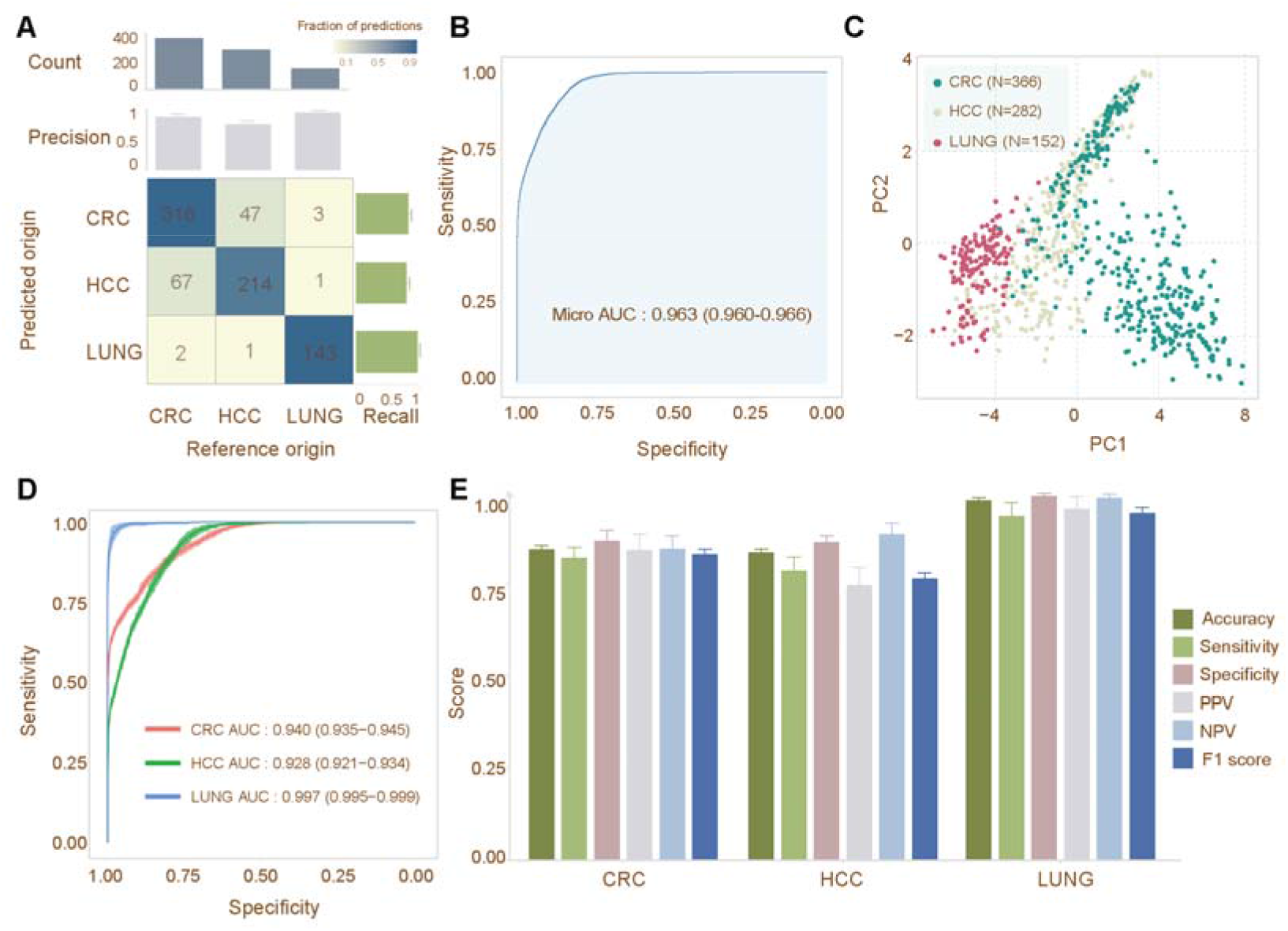
Classification performance of DECIDIA on the detection of different cancer types. **(A)** Confusion matrix and barplot representation of the recall and precision. **(B)** The micro-average ROC curve. **(C)** Scatter plot representation of PCA of learned features. **(D)** ROC curves for the detection of each cancer type in one-versus-rest settting. **(E)** Classification performance for each cancer type.

### Sequence patterns learned by DECIDIA

The self-attention modules in the autoregressive model provided us a convenient way to explore the sequence patterns learned by DECIDIA (See Methods). We constructed four sequence patterns corresponding to the four self-attention heads of DECIDIA (**See Methods**). We found distinctive sequence patterns in cancer patients versus control groups. For example, we observed that all four self-attention heads are featured by high frequency of G in cancer group while in the control group Heads 0, 2 and 3 are featured by high frequency of C and Head 1 is featured by high frequency of T (**Figure 5A**). In addition, we also observed distinctive sequence patterns among CRC, HCC and lung cancer (**Figure 5B).** Specifically, Head 0 is featured by high frequency of C in HCC and lung cancer and less representation of C but higher frequency of G in CRC. Head 1 is featured by high frequency of G in CRC and HCC. Head 2 is featured by high frequency of A particularly in HCC and lung cancer. Head 3 is featured by combination of A and C in HCC and lung cancer and C, G and T in CRC.

**Figure 5.**
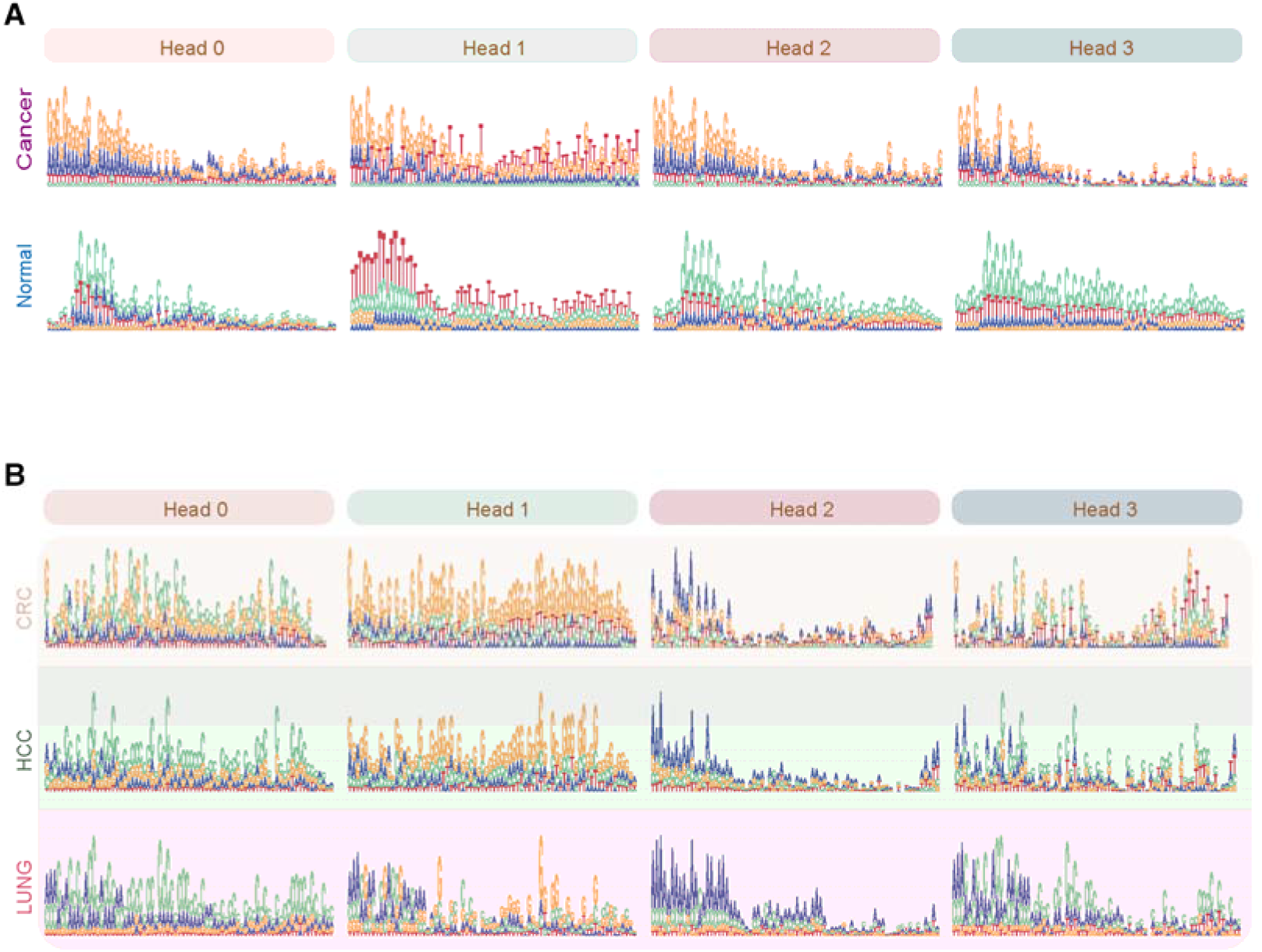
Sequence patterns learned by DECICIA. (A) Four sequence patterns in cancer versus control. (B) Sequence patterns in CRC, HCC and lung cancer.

## Discussion

Accurate early cancer diagnosis is beneficial for improving prognosis.(Shieh et al., 2016) In this study, we proposed a new analytical approach for cancer diagnosis exclusively from the raw sequencing reads obtained from bisulfite sequencing of the plasma cfDNA. We comprehensively evaluated this approach on four datasets for cancer diagnosis and cancer type localization. Although conceptually simple, the proposed approach eliminates the tedious data analytical procedures and is data-efficient and robust.

There are numerous studies exploring the use of bisulfite sequencing of plasma cfDNA for early cancer diagnosis.(Klein et al., 2021; Liu et al., 2020; Luo et al., 2020; Xu et al., 2017) All of these studies require quantitative measurement of methylation as the first step. For example, the methylation value (also known as β-value) for each CpG locus from bisulfite sequencing must be calculated prior to development of machine learning classifier. The β-value is defined as the number of methylated alleles among all alleles at that CpG locus.(Stackpole et al., 2022) However, β-value is reported to be insensitive if the content of tumor-derived cfDNA is low.(Li et al., 2021) To overcome this limitation, Stackpole and colleagues proposed the α-value, defined as the percentage of methylated CpGs among all CpG loci in sequencing reads, to replace the β-value.(Stackpole et al., 2022) However, the calculation of both α-value and β-value requires sequence alignment and postprocessing steps to define methylated and unmethylated cytosine. Sequence alignment of bisulfite sequencing reads is complicated and tedious.(Krueger and Andrews, 2011; Xi and Li, 2009) Bisulfite treatment destroys the complementarity of Watson and Crick strands and leads to asymmetric distribution of C/T in that T in the sequencing reads can be aligned to C in the reference but not vice versa.(Krueger et al., 2012) A common solution is to convert all Cs to Ts for reads and map them to the converted references; subsequently, additional post-processing steps are required to process the alignment results.(Xi and Li, 2009)

DECIDIA achieved at least comparable or better performance as compared with previous studies that developed machine learning classifiers on quantitative cfDNA methylation levels.(Y. Gao et al., 2022; Lin et al., 2021; Ren et al., 2022) In the study conducted by Luo and colleagues, they reported an AUROC of 0.96, sensitivity of 87.9% and specificity of 89.6% in diagnosis of CRC(Luo et al., 2020) with random forest(Breiman, 2001) classifier by using the differential methylation markers. In contrast, DECIDIA achieved an AUROC of 0.98, sensitivity of 0.972 and specificity of 0.911, underscoring the high classification performance.

DECIDIA is data efficient. We demonstrated this advantage with varying number of samples and sequencing reads ranged from two hundred to one thousand with a step size of two hundreds. We observed that the performance of DECIDIA on the minimum data size versus maximum data size are marginal. DECIDIA achieved an AUROC of 0.939 when it was developed with 200 samples and 200 reads and an AUROC of 0.980 when it was developed with 1000 samples and 1000 reads (**Figure 2E and Supplementary Table S2**).

The performance of DECIDIA is robust. This is exemplified by the high performance of our approach on the independent HCC testing set. Although the HCC testing cohort was not involved in the model development, DECIDIA achieved an AUROC of 0.910 on this HCC testing set. This result suggested that there is shared methylated cfDNA patterns among different cancer types and DECIDIA succeeded to learn these shared features. We indeed observed similar patterns of methylated cfDNA from the CRC and HCC datasets (**Figure 5B**).

The use of raw sequencing reads confers several advantages over previous studies that relies on the quantitative cfDNA methylation abundance. Firstly, our solution might be more sensitive towards samples with extremely low content of plasma cfDNA in that precise quantification of tumor methylation levels from low content cfDNA is error-prone.(Q. Gao et al., 2022) Secondly, the methylation signatures of a single cfDNA fragment might be insensitive towards batch effect as compared with quantitative cfDNA methylation abundance; Thirdly, our approach is intuitively and conceptually simple as it does not require base conversion of the sequencing reads and sequence alignment, leading to faster development of diagnostic model and prediction once deployed. In addition, our approach can use as diverse as DNA fragments from the complete cfDNA repertoire whereas alignment-based approaches may discard the exogeneous DNA fragments if they cannot be mapped onto the human genome reference. The exogeneous DNA fragments deriving from bacteria and virus have been demonstrated to be helpful for cancer detection.(Dohlman et al., 2022; Narunsky-Haziza et al., 2022) These merits of our approach would revolutionize the way of plasma cfDNA methylation being used in cancer diagnosis.

However, our approach was not without limitations. The DNA sequence representation learning model was developed exclusively with the human reference genome but not bisulfite sequencing reads. Given that bisulfite treatment changes the unmethylated cytosine to thymine, the sequence patterns of methylated DNA fragments from bisulfite sequencing would be different the reference genome sequence; therefore, features of bisulfite sequencing reads learned by the model developed with human reference genome would be suboptimal. The performance of our approach would be boosted if the feature representation model of DNA sequences were developed with the raw bisulfite sequencing reads. In addition, the weakly-supervised learning model used for classification does not model the inter-dependence among different reads but treat them independently. Improvement of the algorithm allowing it to automatically model inter-dependence of sequencing reads has the potential to increase the performance of the diagnostic model. We will investigate the aforementioned limitations in future studies. In addition, the TNM staging information is not available from these published studies(Hao et al., 2017; Luo et al., 2020; Xu et al., 2017) where the sequencing data were collected. Therefore, so we are not able to perform subgroup analysis in terms of TNM stages. Besides, the number of cancer types used for evaluation of cancer type localization are limited given that the freely available raw sequencing data of plasma cfDNA subjected to bisulfite sequencing is still limited.

Apart from the discrimination between cancer patients from control individuals, our approach is able to identify sequencing reads that are most relevant for classification. Analyses of the most relevant reads lead us to identify distinctive motifs in cancer versus control groups and analogous motifs among different cancer types.

The diagnostic model developed in our study is light-weighted in that it can be run locally on the web browser. We set up a website to provide free access to the developed model for testing and benchmarking purpose. The website is freely available at http://lixiangchun.github.io/DECIDIA/index.html.

In conclusion, we herein reported a deep learning approach for early cancer diagnosis exclusively from the raw sequencing reads of plasma cfDNA subjected to bisulfite sequencing. DECIDIA eliminates the complicated and tedious data analytical procedures. DECIDIA represents a new paradigm of diagnostic model development for liquid biopsy.

## Methods

### Data collection and preprocessing

We collected the sequencing data of 5389 individuals that were subjected to bisulfite sequencing of plasma cfDNA from The Sequence Read Archive (SRA) database. The Accession Numbers are PRJNA574555(Luo et al., 2020), PRJNA360288(Xu et al., 2017), PRJNA383358(Hao et al., 2017) and PRJNA383370(Hao et al., 2017). Detailed information for these datasets is provided in Supplementary Table 1. PRJNA574555 contains 801 patients with colorectal cancer (CRC) and 1021 healthy individuals. PRJNA360288 consists of 1181 patients with hepatocellular carcinoma (HCC) and 959 healthy controls. PRJNA383358 includes 773 patients with CRC. PRJNA383370 has 654 patients with lung cancer. We randomly obtained 1000 reads from each individual. We dropped reads with ambiguous base or average base quality score < 30.

### DECIDIA for cancer diagnosis

We used the CRC dataset (PRJNA574555) for developing cancer diagnostic model. We adopted the 10-fold Monte Carlo cross-validation. Within each cross-validated fold, we randomly partitioned the CRC dataset into a training set (n = 1000), validation set (n = 200) and testing set (n = 622). We used the training set to train the model and validation set to choose the best checkpoint and subsequently evaluated its performance on the testing set. In addition, we used the HCC dataset (PRJNA360288) as an external testing set.

### DECIDIA for cancer type detection

We curated a dataset of 3409 cancer samples from PRJNA574555, PRJNA360288, PRJNA383358 and PRJNA383370 including colorectal cancer (CRC, n = 1154), HCC (n = 1181) and lung cancer (n = 654). We adopted 10-fold Monte Carlo cross-validation. Within each cross-validated fold, we split this dataset into training (n = 2308), validation (n = 300) and testing (n = 800) sets. We used the training set to train the model and validation set to choose the best checkpoint and subsequently evaluated its performance on the testing set.

### Preprocessing of DNA sequence

We constructed a dictionary ***D*** with seven tokens, i.e., ***D*** = {<*s*>, <*e*>, <*pad*>, A, C, G, T}. According to previous study, we added <*s*> and <*e*> to the beginning and end of the input DNA sequence, respectively. The <*pad*> token is used to pad the input DNA sequence to a fixed length if its length is less than a predefined value. The input DNA sequence is truncated if its length exceeds the predefined value. In this study, we set the predefined DNA sequence length to 75. For an input DNA sequence ***S***, e.g., ***S*** = GAGTACG, it was first converted into a character vector ***B*** = {<*s*>, G, A, G, T, A, C, G, <*e*>}, subsequently it was converted into the integer vector of indices according to the dictionary ***D***, i.e., ***X*** = {0, 5, 3, 5, 5, 3, 4, 5, 1}. ***X*** is the input fed to DECIDIA.

### Feature representation learning of DNA sequence

We used the autoregressive language model to learn the feature representation of DNA sequences. Specifically, for an input DNA sequence of length *n* bases ***S*** = {*b*_1_, *b*_2_, *b*_3_ …, *b_n_*}, the purpose of autoregressive modeling is to predict the base *b_i_* in the context of all its preceding bases, i.e., *b*_1_, *b*_2_, *b*_3_ …, *b_i_*_-1_. The standard language modeling objective *L*(***S***) = ∑*_i_ log P*(*b_i_*|*b*_1_, *b*_2_, *b*_3_ …, *b_i_*_-1_; θ) is used to maximize the likelihood. ***θ*** are the parameters of the autoregressive model that is used to model the conditional probability. We used DNA sequences clipped equally from the human reference genome. In our study, we used a single-layer transformer decoder for modeling the “language grammar” of DNA sequence. This language model employs a transformer block followed by softmax to output the distribution over each nucleotide. The transformer block consists of multi-headed self-attention operation and position-wise feed-forward layer. Let *W_e_* denote the nucleotides embedding matrix and *W_p_* the position embedding matrix. This language model can be formulated as(Radford et al., n.d.):

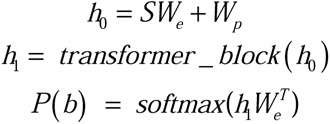

The self-attention head *i* applies the scale dot-product attention to map a query *Q_i_* and a set of key-value pairs ( *K_i_* and *V_i_* ) to output. *Q_i_*, *K_i_* and *V_i_* are projections from the input embedding layer that encodes the input DNA sequence. Let *d_k_* be the dimension of query and key values. Self-attention is defined as:

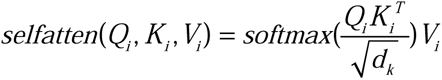

Multi-headed self-attention is simply a concatenation of multiple self-attention heads:

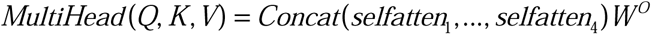

Where *W^O^* is the projection parameter. Position-wise feed-forward layer is defined as:

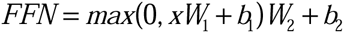

Where *W*_1_ and *W*_2_ are weight matrices and *b*_1_ and *b*_2_ are the bias.

### Attention-based multiple-instance classifier

In the setting of multiple instance learning, a sample is considered as a bag and reads of that sample are instances. For a given sample with *k* reads, we can obtain a feature matrix, denoted as:

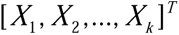

*X_i_* is the feature for the *i*^th^ read outputted from the DNA sequence language model.

A two-layer fully connected neural network is used to transformed the learned DNA sequence feature into latent vector *h_i_*.

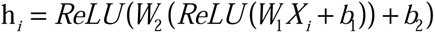

*W*_1_, *W*_2_, *b*_1_ and *b*_2_ are parameters and *ReLU* is the activation function. The attention weight *a_i_* for *h_i_* is defined as(Ilse et al., 2018):

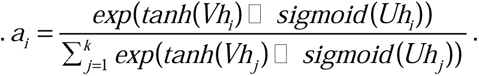

where *V* and *U* are weight parameters; *tanh* and *sigmoid* are activation functions. Attention pooling was applied to obtain the sample-level features:

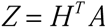

where *A* ={*a*_1_, *a*_2_, *a*_3_…, *a_k_* }, *H* ={*h*_1_, *h*_2_, *h*_3_ …, *h_k_* }. A fully connected layer parameterized as *W*_3_ and *b*_3_ followed by softmax was used to transform the sample-level features into probabilities:

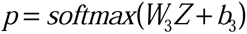

### Construction of sequence patterns

The last row of each attention matrix (denote as *a_i_*) represents the association of each nucleotide in the input DNA sequence on the representation of the input sequence. For each sample, we retrieved the top 100 reads according to the important scores derived from the aforementioned multiple instance classifier. We respectively summed up *a_i_* for reads belonging to different phenotypes stratified by A, C, G and T nucleotides at different positions.

### Training scheme

Feature representation model was pretrained with a batch-size of 4096 for 40 epochs. We used AdamW(Loshchilov and Hutter, 2019) with β1 = 0.9, β2 = 0.999, weight decay of 0.01 and an initial learning rate of 1e-4. The learning rate is warmed up for 3 epochs, and then decays to 0 following a cosine schedule(Loshchilov and Hutter, 2017). The weakly supervised classifier was trained with a batch-size of 1 sample for 100 epochs. We use the cross-entropy loss as the objective function and AdamW optimizer with β1 = 0.9, β2 = 0.999 and weight decay of 1e-5. Learning rate was set to a constant value of 2e-5. All models were trained with PyTorch (version 1.7.1) and transformers (version 4.10.0) on NVIDIA DGX A100.

### Benchmark methods

Motif diversity score(MDS)(Jiang et al., 2020) and end-motif profile deconvolution(Zhou et al., 2023) are two recently proposed statistical methods that characterize cfDNA-derived fragmentome. They have been reported to be applicable for early cancer diagnosis. MDS described the distribution of frequencies of cfDNA end-motif (256 motifs). It is defined as:

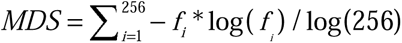

where *f_i_* is the frequencies of end-motif. A higher MDS score indicates a higher fragment diversity.

End-motif profile deconvolution uses non-negative matrix factorization (NMF) to factorize the input end-motif matrix *M* into two non-negative matrices *W* and *F*. Rows of *M* are samples and columns are the frequency of 4-kmer end-motifs. *M* is factorized into *W* and *F* according to:

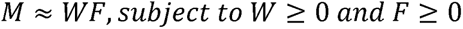

Matrix *W* is the fragmentomic signatures and *F* is referred to F-profile in the study conducted by Zhou and colleagues (Zhou et al., 2023). The F-profile matrix can be used to perform cancer diagnosis.

We directly used the AUC value obtained from machine learning model based on methylation profiles conducted by Luo et al(Luo et al., 2020) as a benchmarked result

## Statistical analysis and software

We used ROC curve, accuracy, sensitivity, specificity, positive predictive value (PPV), and negative predictive value (NPV) to measure the performance of DECIDIA. The ROC curve was created by plotting sensitivity against specificity. The 95% confidence intervals for accuracy, sensitivity, specificity, PPV, and NPV were calculated by Clopper–Pearson method(Julious, 2005). We plotted the ROC curve and calculated AUC with R package pROC (version 1.18.0) and multiROC (version 1.1.1). Statistical analysis was conducted with R software (version 4.1.0), caret package (version 6.0-90) and NMF package (version 0.24.0).

## Ethics statement

This study was approved by the Institutional Review Board (IRB) of Tianjin Medical University Cancer Institute and Hospital and conducted in accordance with Declaration of Helsinki. Informed consent from patients is not applicable and exempted as this study use publicly available data.

## Key Points

- DECIDIA is a deep-learning-based approach for early cancer diagnosis exclusively from raw cfDNA sequencing fragments.
- DECIDIA achieves high classification performance on the internal- and external-testing sets.
- DECIDIA learns distinct fragment signatures directly from raw sequencing reads.
- DECIDIA represents a new paradigm for cancer detection from liquid biopsy.

## Data Availability

The authors declare that the data supporting the findings of this study are available within the paper and its supplementary information files. Raw sequencing data was downloaded from Sequence Read Archive (SRA) with the Accession Number PRJNA574555(Luo et al., 2020), PRJNA360288(Xu et al., 2017), PRJNA383358(Hao et al., 2017) and PRJNA383370(Hao et al., 2017).

## Author Contribution

Xiangchun Li, and Kexin Chen conceived the plan for the project. Xiangchun Li and Jilei Liu designed and performed the experiment. Jilei Liu, Hongru Shen and Xiangchun Li plotted the figures, and wrote the manuscript. Jiani Hu, Xilin Shen, and Chao Zhang collected the data. Yichen Yang, Meng Yang, Wei Wang and Yang Li reviewed the manuscript and figures. Qiang Zhang and Jilong Yang helped in editing the manuscript.

