## Supplementary Figure 1;Supplementary Table 1-4 for "A deep learning approach for cancer diagnosis exclusively from raw sequencing fragments of bisulfite-treated plasma cell-free DNA"

**Supplementary materials**

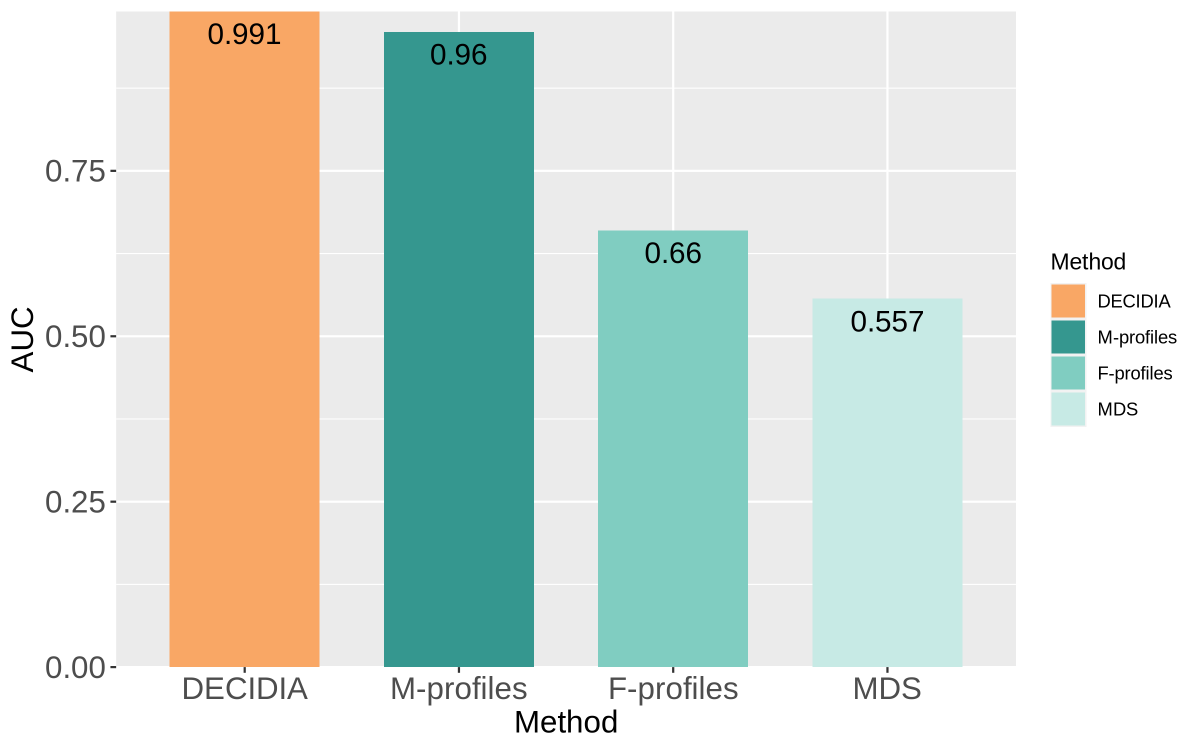

**Supplementary Figure 1. Barplot of AUC values for DECIDIA, M-profile, F-profile and MDS. The AUC value of M-profiles is obtained from study conducted by Zhou and collegues**(Luo et al., 2020).

**Supplementary Table 1. Data source information**

| SRA accession ID | Total samples | Cancer samples | Control samples | Reference | Description |
| --- | --- | --- | --- | --- | --- |
| PRJNA574555 | 1822 | 801 | 1021 | http://www.ncbi.nlm.nih.gov/bioproject/574555 | cfDNA methylation for screening and surveillance of colorectal cancer |
| PRJNA360288 | 2140 | 1181 | 959 | https://www.ncbi.nlm.nih.gov/bioproject/?term=PRJNA360288 | ctDNA methylation markers in diagnosis and prognosis of hepatocellular carcinoma |
| PRJNA383358 | 773 | 773 | NA | https://www.ncbi.nlm.nih.gov/bioproject/?term=PRJNA383358 | Cell-free DNA methylation markers in diagnosis and prognosis of common cancers (COAD) |
| PRJNA383370 | 654 | 654 | NA | https://www.ncbi.nlm.nih.gov/bioproject/?term=PRJNA383370 | Cell-free DNA methylation markers in diagnosis and prognosis of common cancers (lung cancer) |

**Supplementary Table 2. Classification metrics for different number of samples and sequencing reads**

| Number of reads per sample | | Number of samples | Accuracy | Sensitivity | Specificity | Positive predictive value | Negative predictive value | F1-score |
| --- | --- | --- | --- | --- | --- | --- | --- | --- |
| 1000 | 1000 | | 0.939 ± 0.0047 | 0.967 ± 0.0080 | 0.916 ± 0.0118 | 0.903 ± 0.0120 | 0.972 ± 0.0069 | 0.934 ± 0.0044 |
| 1000 | 200 | | 0.907 ± 0.0203 | 0.941 ± 0.0189 | 0.880 ± 0.0253 | 0.863 ± 0.0290 | 0.949 ± 0.0160 | 0.900 ± 0.0222 |
| 1000 | 400 | | 0.934 ± 0.0049 | 0.966 ± 0.0106 | 0.909 ± 0.0150 | 0.895 ± 0.0153 | 0.971 ± 0.0090 | 0.929 ± 0.0048 |
| 1000 | 600 | | 0.933 ± 0.0105 | 0.962 ± 0.0181 | 0.909 ± 0.0109 | 0.895 ± 0.0143 | 0.968 ± 0.0138 | 0.927 ± 0.0130 |
| 1000 | 800 | | 0.933 ± 0.0093 | 0.961 ± 0.0138 | 0.911 ± 0.0168 | 0.897 ± 0.0157 | 0.966 ± 0.0120 | 0.927 ± 0.0088 |
| 200 | 1000 | | 0.936 ± 0.0022 | 0.973 ± 0.0055 | 0.907 ± 0.0054 | 0.893 ± 0.0069 | 0.976 ± 0.0049 | 0.931 ± 0.0029 |
| 200 | 200 | | 0.899 ± 0.0202 | 0.944 ± 0.0239 | 0.862 ± 0.0243 | 0.847 ± 0.0233 | 0.950 ± 0.0211 | 0.893 ± 0.0210 |
| 200 | 400 | | 0.929 ± 0.0042 | 0.962 ± 0.0053 | 0.903 ± 0.0077 | 0.888 ± 0.0068 | 0.967 ± 0.0041 | 0.923 ± 0.0044 |
| 200 | 600 | | 0.936 ± 0.0035 | 0.969 ± 0.0116 | 0.909 ± 0.0117 | 0.896 ± 0.0102 | 0.974 ± 0.0094 | 0.931 ± 0.0033 |
| 200 | 800 | | 0.936 ± 0.0042 | 0.974 ± 0.0076 | 0.905 ± 0.0106 | 0.892 ± 0.0097 | 0.977 ± 0.0061 | 0.931 ± 0.0038 |
| 400 | 1000 | | 0.940 ± 0.0026 | 0.963 ± 0.0091 | 0.921 ± 0.0110 | 0.908 ± 0.0103 | 0.969 ± 0.0072 | 0.935 ± 0.0020 |
| 400 | 200 | | 0.905 ± 0.0203 | 0.935 ± 0.0245 | 0.881 ± 0.0202 | 0.864 ± 0.0232 | 0.944 ± 0.0214 | 0.898 ± 0.0220 |
| 400 | 400 | | 0.932 ± 0.0047 | 0.969 ± 0.0087 | 0.902 ± 0.0112 | 0.888 ± 0.0119 | 0.973 ± 0.0070 | 0.927 ± 0.0054 |
| 400 | 600 | | 0.932 ± 0.0095 | 0.962 ± 0.0191 | 0.908 ± 0.0097 | 0.893 ± 0.0114 | 0.968 ± 0.0142 | 0.926 ± 0.0121 |
| 400 | 800 | | 0.938 ± 0.0041 | 0.969 ± 0.0096 | 0.914 ± 0.0124 | 0.901 ± 0.0129 | 0.974 ± 0.0075 | 0.933 ± 0.0040 |
| 600 | 1000 | | 0.938 ± 0.0043 | 0.972 ± 0.0091 | 0.911 ± 0.0123 | 0.898 ± 0.0114 | 0.976 ± 0.0076 | 0.933 ± 0.0038 |
| 600 | 200 | | 0.897 ± 0.0219 | 0.914 ± 0.0290 | 0.884 ± 0.0195 | 0.863 ± 0.0263 | 0.929 ± 0.0223 | 0.888 ± 0.0258 |
| 600 | 400 | | 0.927 ± 0.0109 | 0.965 ± 0.0117 | 0.898 ± 0.0175 | 0.883 ± 0.0203 | 0.970 ± 0.0095 | 0.922 ± 0.0124 |
| 600 | 600 | | 0.939 ± 0.0063 | 0.971 ± 0.0047 | 0.913 ± 0.0131 | 0.900 ± 0.0137 | 0.975 ± 0.0043 | 0.934 ± 0.0062 |
| 600 | 800 | | 0.934 ± 0.0093 | 0.961 ± 0.0247 | 0.911 ± 0.0098 | 0.897 ± 0.0107 | 0.968 ± 0.0173 | 0.927 ± 0.0127 |
| 800 | 1000 | | 0.940 ± 0.0045 | 0.968 ± 0.0093 | 0.917 ± 0.0119 | 0.904 ± 0.0115 | 0.973 ± 0.0072 | 0.935 ± 0.0043 |
| 800 | 200 | | 0.907 ± 0.0163 | 0.937 ± 0.0258 | 0.883 ± 0.0135 | 0.865 ± 0.0187 | 0.947 ± 0.0195 | 0.899 ± 0.0201 |
| 800 | 400 | | 0.922 ± 0.0137 | 0.952 ± 0.0196 | 0.898 ± 0.0166 | 0.883 ± 0.0174 | 0.959 ± 0.0170 | 0.916 ± 0.0144 |
| 800 | 600 | | 0.938 ± 0.0061 | 0.965 ± 0.0096 | 0.916 ± 0.0135 | 0.903 ± 0.0142 | 0.970 ± 0.0085 | 0.933 ± 0.0059 |
| 800 | 800 | | 0.940 ± 0.0045 | 0.964 ± 0.0125 | 0.920 ± 0.0150 | 0.907 ± 0.0148 | 0.969 ± 0.0104 | 0.934 ± 0.0040 |

**Values are mean ± 95%CI.*

**Supplementary Table 3. Overall accuracy for multi-cancer classification in ten-fold cross-validation setting**

| Fold | Overall accuracy |
| --- | --- |
| 0 | 0.815 (95%CI, 0.786-0.841) |
| 1 | 0.841 (95%CI, 0.814-0.866) |
| 2 | 0.826 (95%CI, 0.798-0.852) |
| 3 | 0.819 (95%CI, 0.790-0.845) |
| 4 | 0.834 (95%CI, 0.806-0.859) |
| 5 | 0.836 (95%CI, 0.809-0.861) |
| 6 | 0.838 (95%CI, 0.810-0.862) |
| 7 | 0.833 (95%CI, 0.805-0.858) |
| 8 | 0.810 (95%CI, 0.781-0.837) |
| 9 | 0.831 (95%CI, 0.803-0.857) |

**Supplementary Table 4. Classification metrics stratified by cancer types**

| Group | Accuracy | Sensitivity | Specificity | Positive predictive value | Negative predictive value | F1-score |
| --- | --- | --- | --- | --- | --- | --- |
| Colorectal cancer | 0.843 ± 0.0074 | 0.821 ± 0.0208 | 0.866 ± 0.0213 | 0.841 ± 0.0313 | 0.845 ± 0.0259 | 0.830 ± 0.0104 |
| Hepatocellular carcinoma | 0.835 ± 0.0067 | 0.786 ± 0.0266 | 0.863 ± 0.0130 | 0.745 ± 0.0352 | 0.884 ± 0.0223 | 0.763 ± 0.0118 |
| Lung cancer | 0.978 ± 0.0048 | 0.934 ± 0.0277 | 0.990 ± 0.0055 | 0.955 ± 0.0245 | 0.984 ± 0.0078 | 0.943 ± 0.0113 |

**Values are mean ± 95%CI.*
